# Parkinson’s disease patients display a DNA damage signature in blood that is predictive of disease progression

**DOI:** 10.1101/2024.03.25.24301713

**Authors:** Daisy Sproviero, César Payán-Gómez, Chiara Milanese, Shixiang Sun, Akos Gyenis, Domenico Delia, Tammaryn Lashley, Jan Vijg, Jan H.J. Hoeijmakers, Pier G. Mastroberardino

**Affiliations:** IFOM-ETS, The AIRC Institute of Molecular Oncology, Milan 20139, Italy; Universidad Nacional de Colombia, Sede de La Paz, La Paz 111321, Colombia; Department of Genetics, Albert Einstein College of Medicine, Bronx 10461, USA; University of Cologne, Faculty of Medicine, Cluster of Excellence for Aging Research, Institute for Genome Stability in Ageing and Disease, Cologne, Germany; The Queen Square Brain Bank for Neurological Disorders, Department of Clinical and Movement Neuroscience, UCL Queen Square Institute of Neurology, London, UK; Department of Neurodegenerative diseases, UCL Queen Square Institute of Neurology, London, UK; Erasmus MC, Rotterdam 3000 DR, the Netherlands; Princess Maxima Center for Pediatric Oncology, Oncode Institute, Utrecht, The Netherlands; Center for Single-Cell Omics, School of Public Health, Shanghai Jiao Tong University School of Medicine, Shanghai 200025, China; Universita’ degli Studi dell’Aquila, L’Aquila 67100, Italy

**Author notes:** DS and CPG are first co-authors and equally contributed to the study.

## Abstract

Aging is the main risk factor for Parkinson’s disease (PD), yet our understanding of how age-related mechanisms contribute to PD pathophysiology remains limited. We conducted a longitudinal analysis of the Parkinson’s Progression Markers Initiative cohort to investigate the involvement of DNA damage in PD. Our findings revealed that PD patients exhibit disrupted DNA repair pathways and biased suppression of longer transcripts, indicating the presence of age-related, transcription-stalling DNA damage. Notably, this DNA damage signature was only detected in patients with more severe motor symptom progression over a three-year period, suggesting its potential as a predictor of disease severity. We further validated this signature in independent PD cohorts and confirmed increased signs of DNA damage in dopamine neurons of the substantia nigra pars compacta through histopathological analysis of PD brains. Our study sheds light on an aging-related mechanism in PD pathogenesis and identifies markers of disease progression providing a readily applicable diagnostic platform to prognosticate disease progression.

**One Sentence Summary:** Parkinson’s disease patients display a DNA damage signature in blood that is predictive of disease progression.

## Introduction

Parkinson’s disease (PD) is a common neurodegenerative disorder affecting principally, but not exclusively, dopaminergic neurons in the nigrostriatal circuits. PD is primarily idiopathic, with less than 10% of cases attributed to monogenic mutations; the genetic forms, however, informed on the intricate pathogenic mechanisms underlying disease progression that include perturbation in protein homeostasis, oxido-reductive balance, mitochondrial function, and intracellular trafficking (*1, 2*). These alterations often interact in a complex manner, resulting in the characteristic motor symptoms observed in PD. The disease, however, is remarkably heterogeneous (*3*) and patients display significant variability from the standpoint of both clinical presentation and progression, which is attributed to differences in the underlying pathophysiological processes (*4*). Consequently, PD has also been considered a syndrome with several disease subtypes (*5, 6*). Our understanding of the causes of PD heterogeneity, however, is extremely rudimentary and stratification of patients in homogeneous groups based on the expected clinical presentation and disease progression remains at present unachievable. This drawback represents a major confounder in clinical trials as treating a heterogeneous cohort of patients is likely to yield varied outcomes (*7, 8*). Conversely, stratification of patients in clinically homogeneous groups would facilitate the development of personalized, more effective treatments. The principal risk factor for PD is aging (*9*). How the biology of aging contributes to PD pathophysiology, however, is poorly understood. A fundamental and causative mechanism of aging is the progressive accumulation of damage in nuclear DNA (*10*). Nuclear DNA is, in fact, constantly exposed to exogenous and endogenous factors that induce chemical modifications of its bases that ultimately corrupt genetic information fidelity. These chemical alterations impact cellular function, for instance causing quantitative and qualitative perturbation of transcription and culminate in transmissible mutations in replicating cells. The impact of DNA damage on transcription is highly relevant for non-dividing, postmitotic neurons. Given the severe consequences of DNA damage accumulation, evolution has equipped all organisms with a highly efficient and intricate network to promptly correct these lesions (*11*).

Nuclear DNA damage can explain most -if not all-hallmarks of aging and has been considered as the ultimate underlying cause of age-related systemic functional loss and disease (*12*). Severe defects in DNA repair systems cause progeroid diseases in humans and mice characterized by the premature appearance of multiple symptoms of aging. On the other hand, natural aging is associated with an increased burden of somatic mutations in virtually all organs and tissues including brain and neurons in particular (*13, 14*), which also points towards increased nuclear DNA damage as its underlying cause.

Impaired DNA repair has also been hypothesized to contribute to the pathogenesis of age-related neurodegenerative diseases (*15*) and multiple evidence points to a role in PD as well. Peripheral fibroblasts from PD patients, in fact, display reduced DNA repair capacity (*16*), mouse models with mild defects in DNA repair display an impairment in the nigrostriatal dopaminergic pathways (*16*), and synucleinopathy is associated with activation of the DNA damage response in mice (*17*). Moreover, meta-analysis available via the PD Gene database (http://www.pdgene.org/) (*18, 19*) indicates that the nucleotide excision repair (NER) gene ERCC8 may be associated with PD (e.g. meta P-value=9.27e-07, metaOR=1.15, 95%CI (1.08-1.21) for the rs11744756 polymorphism). Finally, a role for DNA damage accumulation in PD is also substantiated by evidence demonstrating that certain patients suffering from genetic diseases caused by defects in DNA repair manifest levodopa responsive dopaminergic symptoms (*20, 21*). Collectively, these elements suggest that accumulation of nuclear DNA damage may participate in PD pathogenesis. Evidence to fully prove this hypothesis, however, is inconclusive thus far.

In this study we leveraged on the large Parkinson’s Progression Markers Initiative (PPMI) datasets collection to gather further bioinformatics evidence demonstrating a role for nuclear DNA damage in PD. We evaluated longitudinal expression data in 484 PD patients and 187 controls examined at the intake visit (visit 1), and in 268 and 157 of these patients and healthy subject that were examined in a follow up visit after 36 months (visit 8). This cohort consisted of idiopathic cases (iPD), patients carrying genetic mutations in LRRK2 and GBA, prodromal cases (people without motor signs but at a higher risk of developing PD), and healthy controls.

## Results

### Parkinson’s Disease Patients show dysregulation in DNA repair pathways

We initially conducted quality control analyses to identify potential biases in the library preparation. We analyzed datasets from patients examined at the intake visit (baseline or visit 1) and at a follow-up visit after 36 months (visit 8). The size and gender composition of the experimental groups at the two time points are summarized in the table in Figure S1A.

Quality control revealed homogeneous gene counts distribution and comparable library size in the Salmon files ensuring data integrity. Principal component analysis (PCA) further revealed a compact clustering of data in all experimental groups. In PD genetic and idiopathic patients, 44% of the variance was attributable to PC1 at baseline, while at visit 8 PC1 accounted for 37% of the variance (fig.S1B-C). The consistent distribution observed in the PCA plots between the experimental groups – that is iPD, LRRK2 and GBA mutants, and healthy subjects - aligns with previous reports (*22*).

The age of PD patients and healthy controls was comparable (fig.S1D), and the observed effects are therefore genuinely attributable to disease effects rather differences in age. Prodromal cases, however, were significantly older than controls.

To assess the data quality further, we compared the transcriptome of PD patients at visit 1 and visit 8 and calculated the enrichment factor for common deregulated transcripts. Remarkably, expression profiles at the two time points displayed an overlap 26 fold higher than what expected by chance, substantiating the reliability of the datasets (fig.S1E).

Although differentially expressed genes exhibited modest fold changes, their statistical significance was highly pronounced in both visits, as indicated by the adjusted p-values (fig.S2A-B). The number of differentially expressed genes (DEGs) significantly varied between experimental groups and time point, indicating heterogeneity among different forms of PD (fig.1A, fig.S2C). As a cautionary note, we observe that at visit 8 the size of the LRRK2 group remarkably differed from that of visit 1, being composed of 14 patients, i.e. 10% of visit 1 size. This sharp difference obviously weakens the conclusions on LRRK2 experimental group at visit 8. The same partially applies, albeit to a lesser extent, to the GBA group, which at visit 8 included 28 patients, i.e. 56% of visit 1 size.

**Fig. 1.**
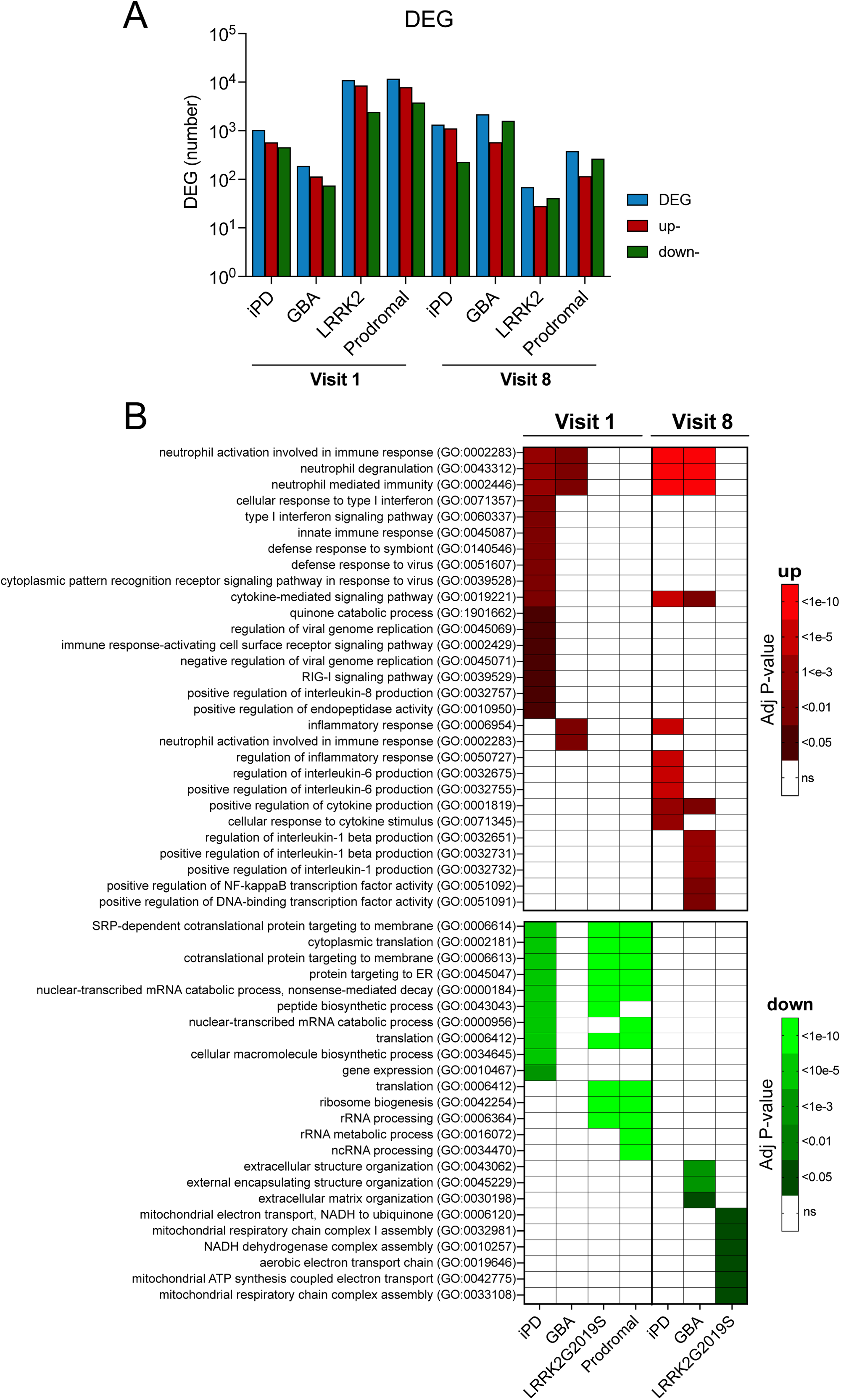
Over-representation analysis of significantly DEG in PD patients’ blood. (**A**) Summary histogram of the number of significantly total, up-, and downregulated DEG (blue, red, and green respectively) in the analyzed experimental groups, at the intake visit and after 36 months (i.e. visit 8). (**B**) Over-represented Gene Ontology up-(red) and down-regulated (green) processes based on significantly DEG at visit 1 and 8. Gray boxes indicate that the process is not altered. Upregulated processes are largely related to inflammatory processes, while the down-regulated ones predominantly concern macromolecular synthesis. The sensitivity of the analysis based on significantly DEG is modest and failed to detect alterations, for instance in GBA patients or prodromal cases at visit 1. In both (**A**) and (**B**) significance was set at adjusted P-value <0.05. Adjusted P-values were obtained using Fisher’s exact test.

To gain further insights into the molecular processes altered in PD and its different forms, we performed pathway analysis using two complementary approaches. First, we conducted a classical overrepresentation analysis (ORA) (*23*), followed by a Gene Set Enrichment Analysis (GSEA) (*24*).

ORA confirmed the heterogeneity between PD forms and between the timepoints of observations. For instance, several upregulated pathways were unique for iPD at visit 1, while others were unique to GBA or LRRK2 at visit 8 (fig.1B). The analysis also revealed that upregulated processes were largely related to inflammation, consistent with previous studies (*22*), while downregulated processes mostly relate to macromolecular synthesis and mitochondrial biology. The downregulation of macromolecular synthesis pathways is noteworthy, considering the well-established involvement of mitochondria in PD etiopathogenesis. ORA failed to identify significantly upregulated processes in LRRK2 and prodromal patients. We note, however, that because ORA processes significantly DEG, the analysis necessarily returns information that is based on relatively pronounced differences. We reasoned, however, that functional differences could stem from small changes in groups of related genes that are coordinately expressed, i.e. expression modules, which cannot be detected by a method bound to significant DEGs. This limitation is addressed by GSEA (*24*), which used all the ranked genes in the datasets according to their individual statistical significance, to calculate aggregated statistical significance in a set of related genes, which constitute a pathway.

GSEA against the highly detailed Reactome database identified a total of 839 deregulated pathways in the four groups (i.e. iPD, GBA, LRRK2, and prodromal), at visit 1, with 471 unique elements. Only 15 pathways, however, were common to the four experimental groups and were all downregulated (fig. 2A, B, S2D). Although these pathways included expected processes such as mitochondrial function, they primarily related to macromolecular synthesis (fig. 2C). Interestingly, the shared pathways at visit 8 were different from those identified at visit 1 and included processes related to immunity, cell death, disease, and senescence, a direct consequence of DNA damage accumulation (*25*) (fig. 2D-F). Expression variations in the LRRK2 group were largely in the opposite direction than those observed in iPD and GBA (fig. S2E), an effect that can likely ascribed to the very small size of this group at visit 8 (N=14).

**Fig. 2.**
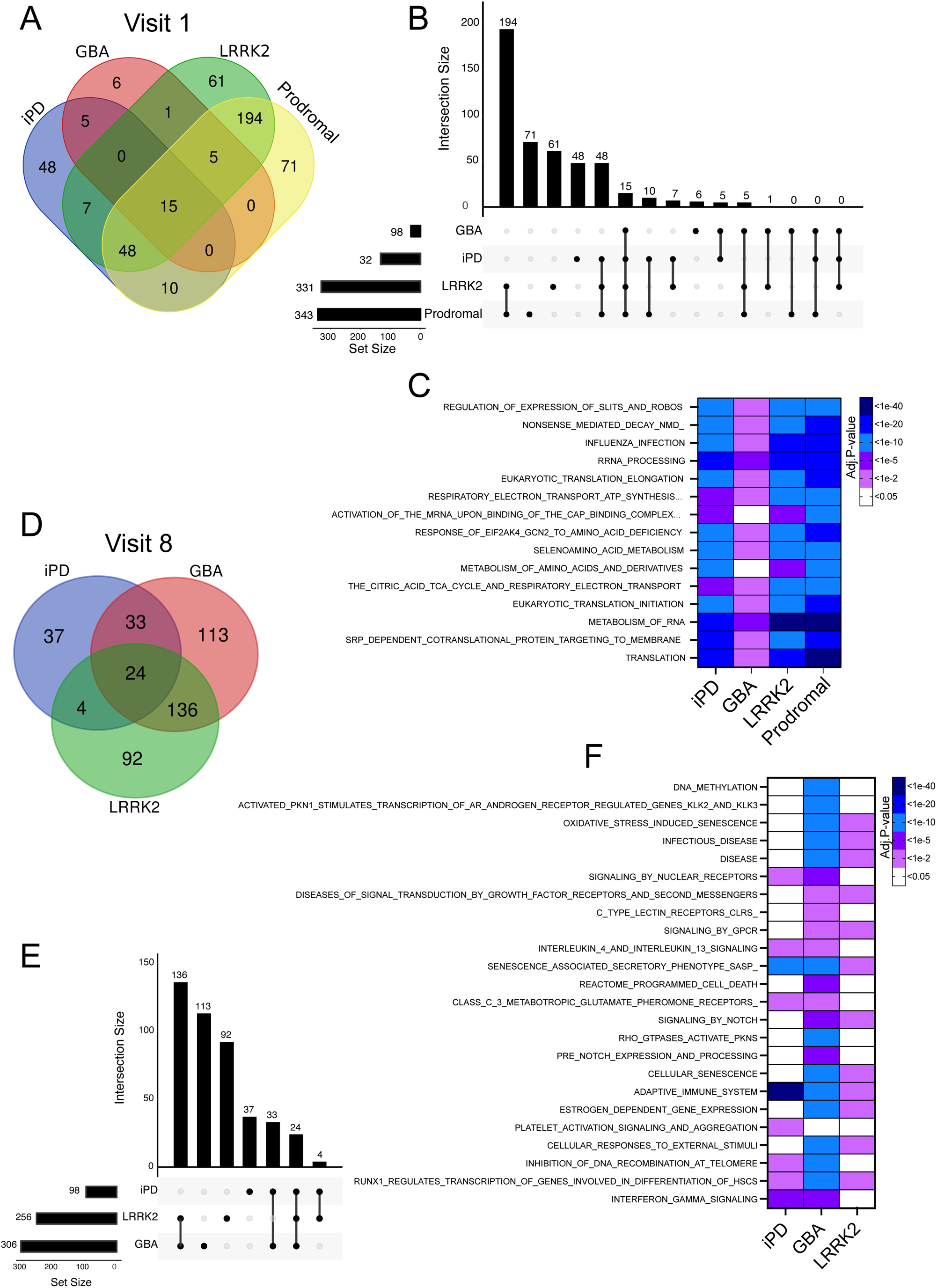
Differentially regulated pathways identified by gene set enrichment analysis (GSEA). (**A**) Venn diagram and (**B**) UpSet plot of enriched GSEA Reactome pathways indicate that 15 are shared among the different analyzed groups. (**C**) The 15 common pathways at visit 1 include mechanisms related to macromolecular synthesis, mitochondrial function, and immunity. (**D**) Venn diagram and (**E**) UpSet plot of enriched GSEA Reactome pathways to display intersections between datasets shows 24 common pathways among the studied groups at visit 8. (**F**) Common pathways at visit 8 differ than those identified at visit 1 and include general disease related processes, senescence, and inflammation.

The observed alterations in macromolecular synthesis may indicate genome instability, as DNA damage is intrinsically associated with transcription and downstream translation (*26, 27*). Moreover, macromolecular synthesis has been hypothesized to be perturbed in PD (*28*), despite extremely rudimentary understanding of its role in the disease, and previous transcriptome analysis revealed alterations in ribosome-related pathways in PD patients’ brains that are also detectable in DNA-repair-defective mice (*16*). To further examine whether macromolecular synthesis is perturbed during early symptomatic PD (i.e. visit 1), we performed transcriptome examination at a more granular level taking advantage of leading-edge gene (LEG) analysis. GSEA, in fact, identifies the subset of genes, i.e. LEG, that provide the highest contribution to the enrichment signal of a given pathway (*24*). LEG therefore reduces the dimensionality of enriched gene sets (i.e. several of the GSEA identified pathways contain the same genes), providing a focused depiction of transcriptomic changes and facilitating the identification of crucial biological processes. We extracted the LEG contributing to the top 20 deregulated pathways in the datasets and asked which biological processes are governed by the 178 common dysregulated genes (fig.3A). The analysis revealed processes largely related to RNA metabolism (fig.3B, C), therefore confirming the results of the previous analysis and supporting the rationale for investigating DNA repair in PD.

**Fig. 3.**
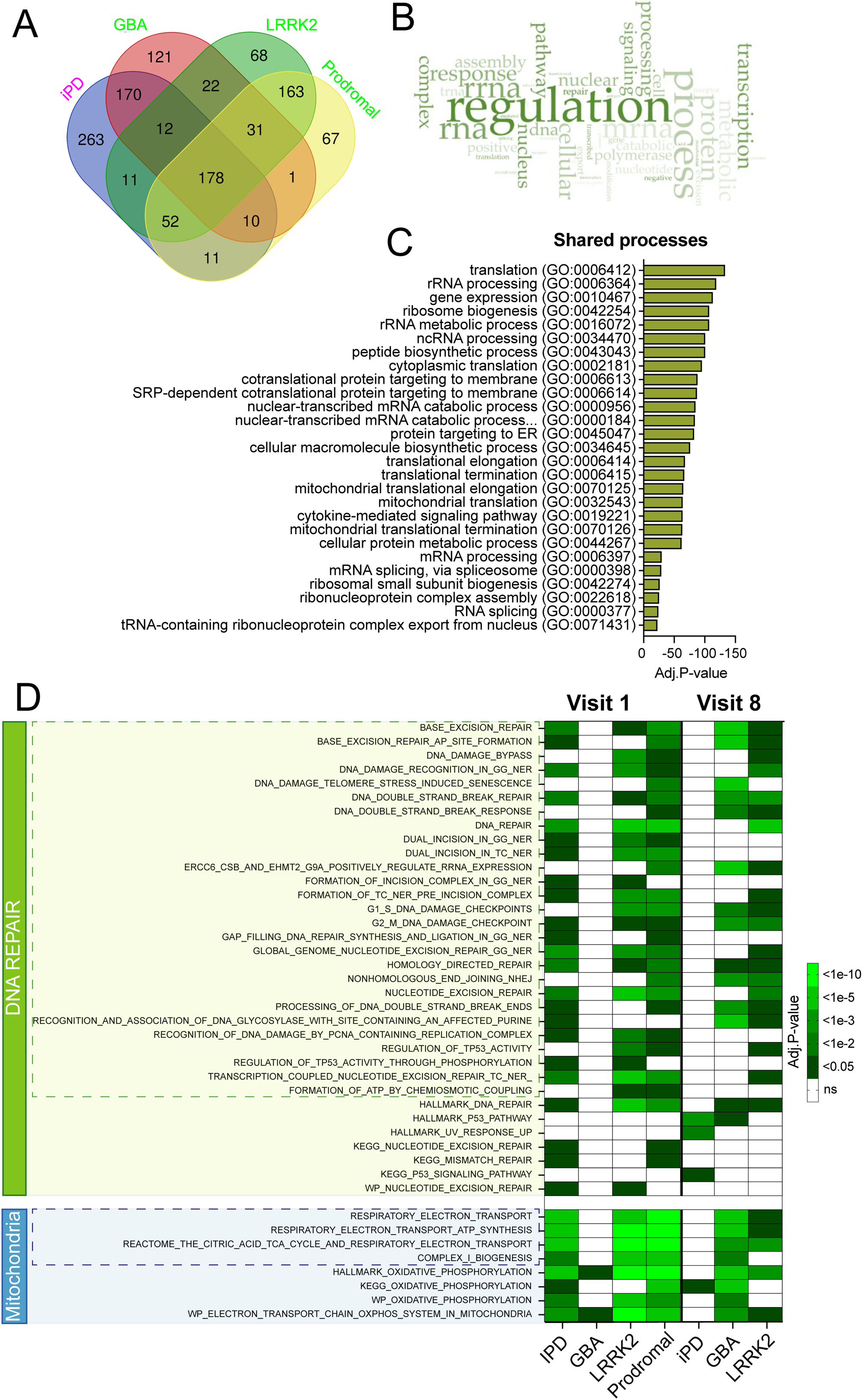
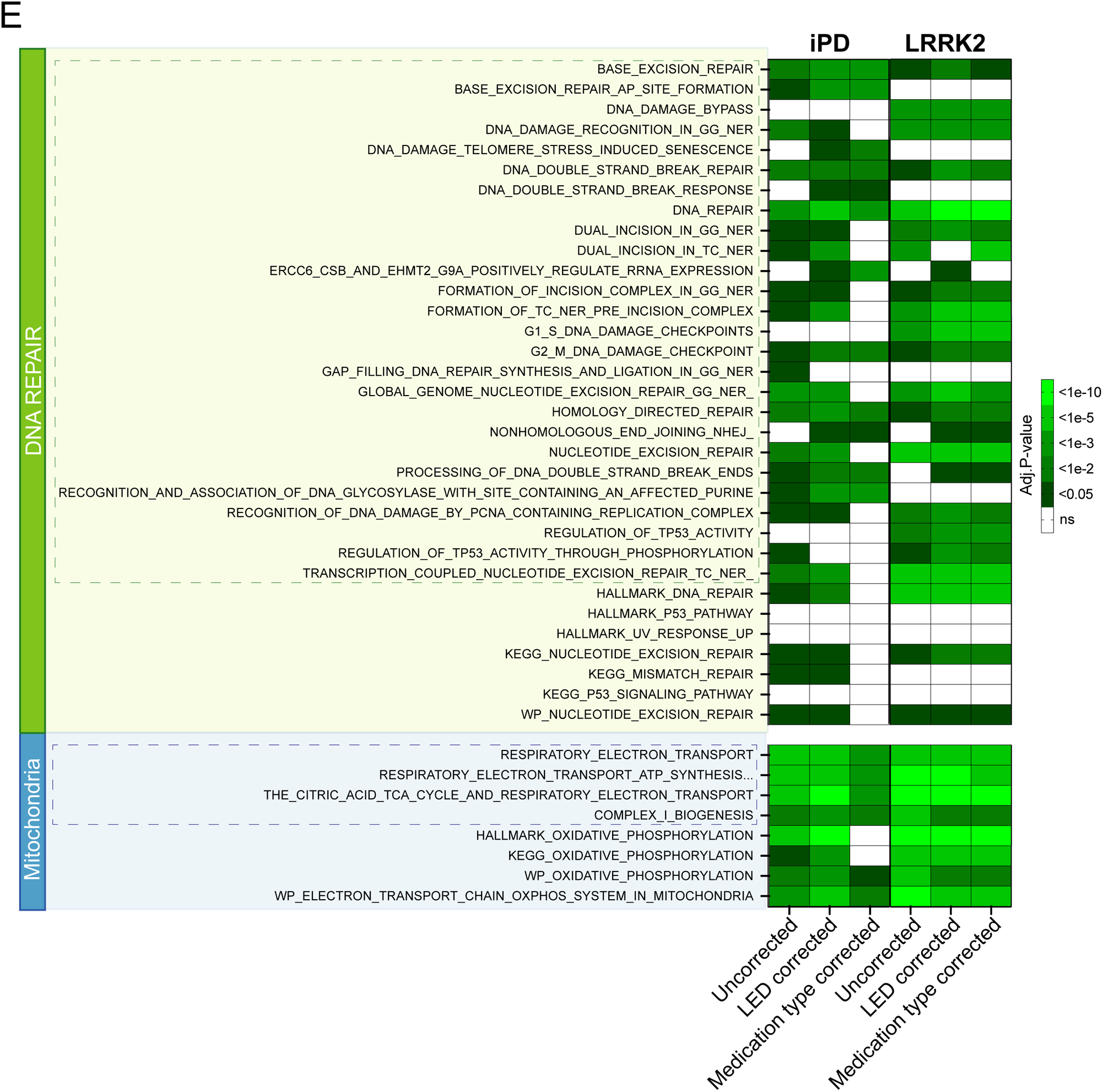
Leading edge gene analysis and GSEA identify alteration in processes related to macromolecular synthesis and DNA repair. **(A)** Venn diagram based on the leading edge genes (LEG) identified by GSEA providing the most significant contribution to the top 20 deregulated pathways in the datasets. 178 genes are common to the studied experimental groups. (**B**) Text mining indicates that pathways related to the 178 LEG related to processes involving RNA, transcription, and polymerase activity, all of which are intertwined with DNA damage and repair. **(C)** Histogram showing the deregulated processes governed by the identified LEG. lea GSEA pro and C) The analysis revealed processes largely related to RNA metabolism. (**D**) Several DNA repair pathways are downregulated in PD, albeit at different time points. GBA patients, for instance, display alterations only at visit 8, while downregulation is prominent at visit 1 in iPD patients. As expected, genes related to mitochondrial function and complex I assembly are downregulated in PD. (**E**) Alterations in DNA repair pathways persist after correction for medication intensity, i.e. daily levodopa equivalent dose (LED) and type of medications in both iPD and G2019S LRRK2 carriers. GBA patients were not analyzed as they did not display alterations in DNA repair pathways at visit 1. Reactome pathways are delimited by dashed boxes.

Subsequently, we directly investigated whether DNA repair pathways are perturbed in PD and determined their up- or down regulation using GSEA. The analysis revealed downregulation of several Reactome DNA repair pathways at visit 1 in iPD, LRRK2, and prodromal cases (fig. 3D). Downregulated pathways included NER and the data are therefore consistent with our previous findings showing deranged NER capacity in iPD and LRRK2 patients (*16*). GBA patients, on the other hand, showed relatively unaffected DNA repair pathways at visit 1 but displayed downregulation in many DNA pathways at visit 8. Interestingly, in the follow up visit, alterations were remarkably reduced in iPD patients, and the only group that continued to exhibit defects was that of LRRK2 cases.

To ensure the reliability of our findings, we also monitored DNA repair pathways using additional platforms, namely KEGG, Hallmarks, and Wikipathways, all of which confirmed our observations in Reactome (fig.3D). Importantly, the analysis also displayed downregulation of mitochondrial pathways (fig.3D), which is expected given the central role of mitochondrial dysfunction in PD. Collectively these results demonstrate that DNA repair mechanisms are reduced in PD, albeit with different kinetics during disease progression among its different forms.

### Alterations in DNA repair pathways persist after correction for medications

To investigate whether the observed changes in DNA repair pathways were influenced by medication, we performed data analysis considering medication intensities, specifically the levodopa equivalent dose (LED) (*29*), and medication types. We included medication variables as covariates in our statistical models. In particular, medications were included in regression analysis as independent variables so that the the model could account for their potential influence on the observed GSEA pathways. Daily LED was treated as a continuous variable. Medication type was treated as a categorical variable and regimens were divided into seven groups: 1) only levodopa/carbidopa, (2) only dopamine agonists, (3) levodopa/carbidopa and dopamine agonists, (4) levodopa/carbidopa and other medications, (5) levodopa/carbidopa, dopamine agonists, and other medications, (6) other medications, (7) no medications. When the medication filed was left blank in the PPMI record, the patient was classified as not taking any medication. No patients were treated with dopamine agonists in combination with other medications. Other medications included A2A receptor antagonists, MAOB inhibitors, COMT inhibitors, and anticholinergics.

Because minimal changes in DNA repair pathways were observed in idiopathic patients at visit 8 (fig. 3D), we focused on visit 1 and therefore on potential predictive early markers of disease progression. GSEA revealed that changes in DNA damage-related pathways were generally unaffected by LED, with statistical significance being lost in only 8 out of the 33 altered pathways in iPD (fig. 3E). However, when correcting for medication type in iPD significance was compromised in 21 out of the 33 pathways. It is important to note that the correction for medication also resulted in the loss of significance in the KEGG and HALLMARK oxidative phosphorylation pathways, which is inconsistent with PD pathogenesis considering the well-established role of mitochondrial dysfunction in the disease and the significant changes observed in other key mitochondrial pathways (fig. 3E). We also note that loss in statistically significant pathways in the correction analysis is consistent with intrinsic reduction of statistical power, which is in turn caused by sample size reduction due to the introduction of additional variables in the analysis.

LRRK2 patients’ data were less affected by medication correction and all but one of the pathways downregulated in the uncorrected analysis (REACTOME_ERCC_CSB…) was not significantly altered after correction (fig.3E). This result may indicate a more profound impact of DNA damage and repair in a G2019S LRRK2 background, which is also consistent with previous data from our laboratory obtained in fibroblasts (*16*). We did not perform any corrected analysis for GBA patients as they did not show any alterations in DNA repair pathways at visit 1 (fig.3D).

Collectively, these data show a modest impact of medication on DNA repair pathways alterations. This result is also consistent with the observation that people with prodromal signs, who should not be taking PD medications, display similar changes in DNA repair pathways (fig. 3D).

LEDD and medication type are highly correlated variables, interacting with each other, and are therefore confounding variables. Given these conditions, including all the variables in the same correction model may lead to issues involving multicollinearity, which hampers accurate evaluation of their individual effect (*30*). For this reason, we did not perform correction analysis simultaneously using both LEDD and medication type.

### Biased reduction of long genes transcription levels in PD points to DNA damage accumulation

DNA damage is considered as a stochastic event, and it is therefore more likely to occur in longer than in shorter genes. Because DNA damage can block RNA polymerase progression and therefore transcription, its accumulation is paralleled by a biased reduction of longer genes’ transcripts. Indeed, our and other laboratories demonstrated that <u>a</u>nalysis for length-<u>b</u>iased <u>a</u>lterations in <u>tr</u>anscription <u>o</u>utput (ALBATRO) mirrors DNA damage accumulation (*26, 31, 32*).

To make ALBATRO informative, a sufficiently large number of DEG must be identified. In this study, we arbitrarily set this number to be at least 100 genes per distribution. In visit 1 specimens, ALBATRO did not reveal any biased reduction in longer gene transcripts when compared to healthy controls: longer genes were even more represented in PD specimens, but the test was not statistically significant (median length up-regulated genes 29753 bp, median length down-regulated genes 19076 bp, p.Wilcoxon=0.0555) (fig. 4A, D). Overrepresentation of longer genes may reflect an adaptive protective response given recent evidence indicating that longer transcripts are often related to anti-aging genes (*33*). At visit 8, however, ALBATRO did detect significant down-regulation of longer-transcripts in PD specimens (median length up-regulated genes 25706 bp, median length down-regulated genes 40692 bp, p.Wilcoxon=0.0011) (fig. 4B, D). Interestingly, also prodromal cases showed biased downregulation of longer transcripts (fig.4C, D).

**Fig. 4.**
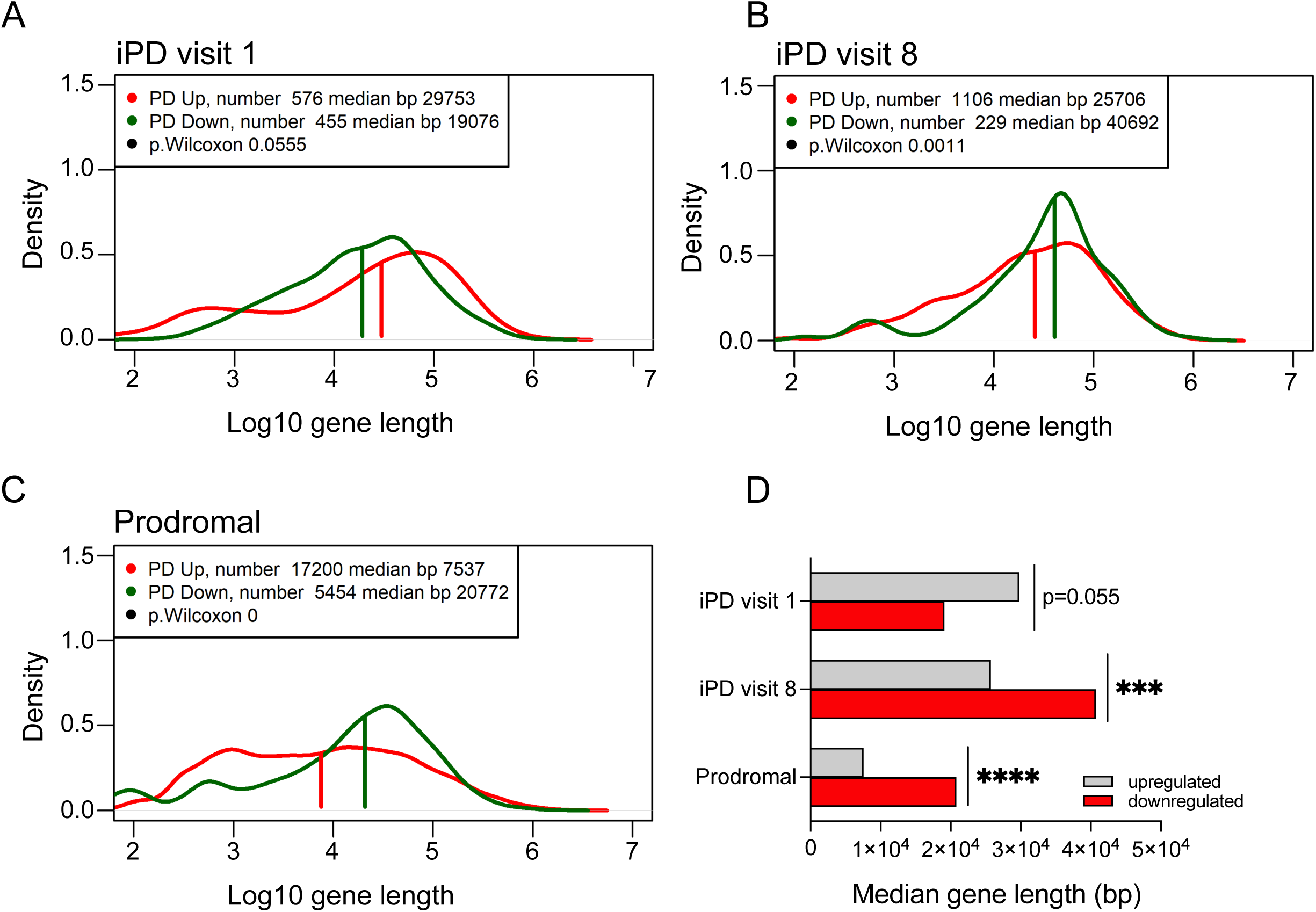
Biased reduction in the expression of longer genes in PD. Frequency plots of gene length of up- and downregulated genes (red and green trace respectively) in iPD at visit 1 (**A**) and at visit 8 (**B**), and in prodromal cases (**C**). Histogram summarizing the findings in frequency plots showing that downregulated genes are significantly longer in iPD at visit 8 and in prodromal cases. P-value for iPD at visit 1 is close to significance (p=0.055). (**D)** Bar graph summarizing the detected effects. Statistical significance of the distributions was tested with Wilcoxon signed-rank test (p.Wilcoxon, p<0.001 ***, p<0.0001 ****).

ALBATRO detected slight effects in genetic PD cases (GBA, LRRK2 G2019S). At visit 1, GBA patients did not display significant differences (data not shown). At visit 8, instead, GBA mutants displayed a statistically significant difference in the length of up-vs. down-regulated genes, albeit in the opposite direction of that associated with DNA damage accumulation, i.e. shorter genes were upregulated (median length up-regulated genes 31529 bp, median length down-regulated genes 17384 bp, p.Wilcoxon<1e-07) (fig.S3A). This evidence points again to different pathogenic mechanisms in GBA mutants. Instead, patients carrying LRKK2 G2019S at visit 1 displayed a downregulation of longer genes compared to shorter, but the test was not statistically significant (median length up-regulated genes 12216 bp, median length down-regulated genes 17417 bp, p.Wilcoxon=0.2509) (fig.S3B). Similarly, no differences were observed in LRRK2 at visit 8. We note, however the lack of observed effect in these patients might be due to of an unexpected peak in very short downregulated genes (approx. 100 bp, fig.S3B) at visit 1, and to lack of statistical power at visit 8 (group size N=14).

Because results from bioinformatic analysis may depend upon the studied dataset, we applied ALBATRO to another available dataset of PD blood transcriptome (GSE99039), composed of 205 iPD and 233 controls (*34*). The analysis returned the same results (fig.S3C), therefore validating our data in the PPMI cohort.

### Transcriptional deregulation of DNA repair pathways predicts disease severity

Our results indicate that down-regulation of DNA repair pathways occurs early in the symptomatic stage of PD (i.e. is detectable at visit 1) and that surrogate measures of DNA damage accumulation (i.e. biased decline of long transcripts) increase with disease progression. We therefore asked whether deregulation of DNA repair pathways may inform on the rate of disease progression. We therefore stratified iPD patients examined at both visit 1 and visit 8 in two groups with different progression rate. One group was composed of patients with increased severity as measured by the UPDRS III score (severe group, deltaUPDRS=UPDRS(visit 8)-UPDRS(visit 1)>0, n=226). Another group was composed of patients who did not display worsening of the UPDRS III score (mild group, deltaUPDRS≤1, n=112). We compared these groups to healthy controls (n=152) (fig.5A). Differentially expressed genes are listed in supplementary table 1. GSEA revealed again substantial heterogeneity and detected only 8 shared pathways between mild and severe cases at the two observed timepoints (fig.5B). Four of the identified processes were again inherent to RNA metabolism, while other pathways were related to disease and inflammation in general terms (fig. 5C).

**Fig. 5.**
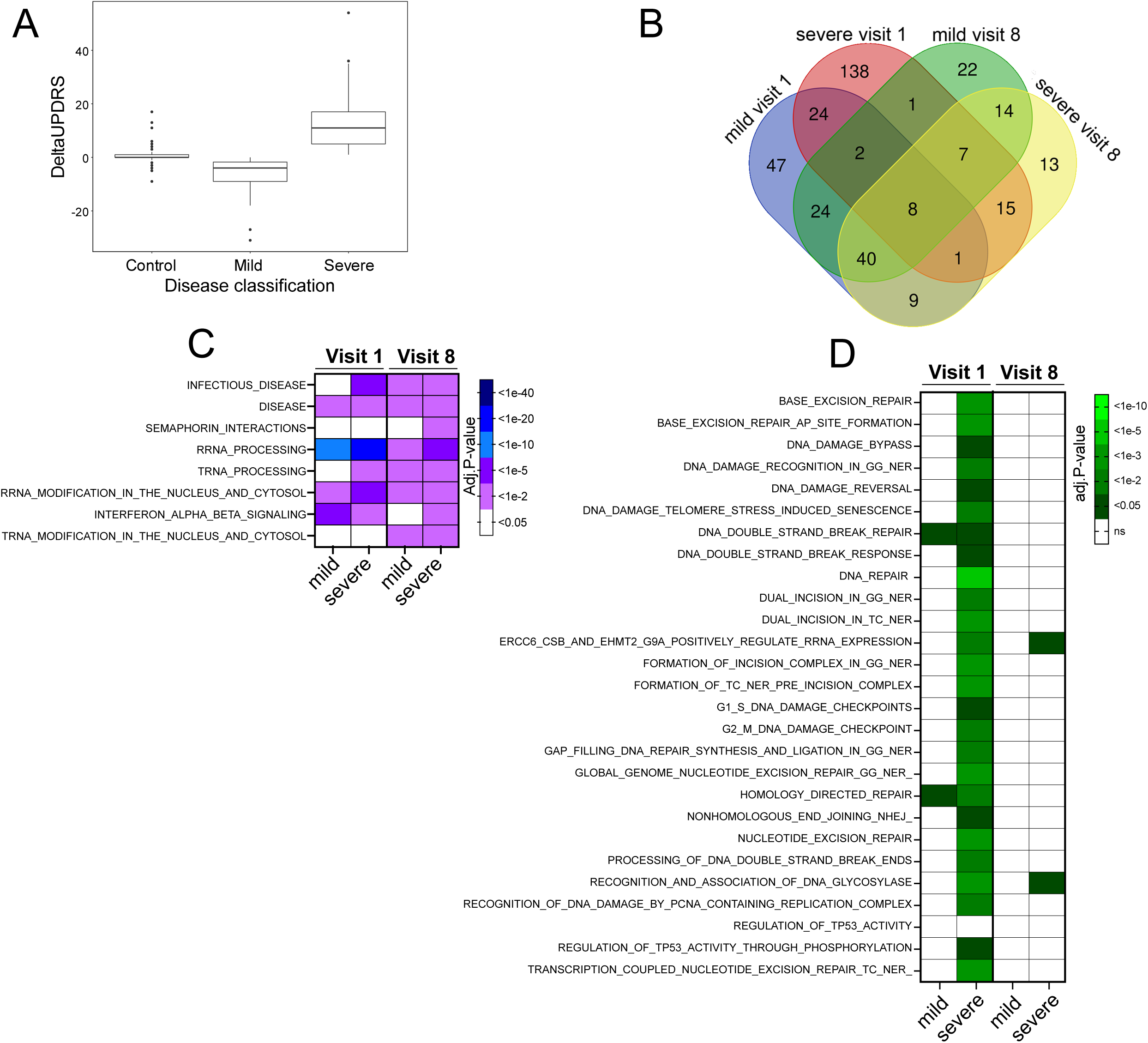
GSEA in patients with different disease progression. (**A**) Summary boxplot of deltaUPDRS, defined as the difference in UPDRS score between visit 8 and visit 1 in iPD patients. GO-BP analysis at visit 1 in mild versus severe patients. (**B**) Venn Diagram of GSEA Reactome pathways detected 14 shared pathways between mild and severe cases at the two observed timepoints, and overall reveals differences between groups and time of analysis. (**C**) The 14 shared pathways relate to general processes of disease and to inflammation. (**D**) DNA repair pathways are predominantly altered in patients with severe progressions, at visit 1.

When we investigated the expression of DNA repair pathways in these experimental groups, we found significant downregulation of many varied mechanisms, only at visit 1, in those patients that displayed deterioration of motor symptoms in follow up visit 8, 36 months later (fig.5D). Consistently with the analysis in unstratified iPD (fig. 3D), we did not detect significant alterations in DNA repair pathways at visit 8.

At visit 1, ALBATRO detected reduced transcription of longer genes only in severe cases; conversely, only mild cases displayed biased transcription reduction at visit 8 (fig.6A, B). Collectively, these elements indicate that measures of defective DNA repair and nuclear DNA damage accumulation may predict severity of disease progression and suggest that different kinetics govern these processes during the symptomatic phase of the disease.

**Fig. 6.**
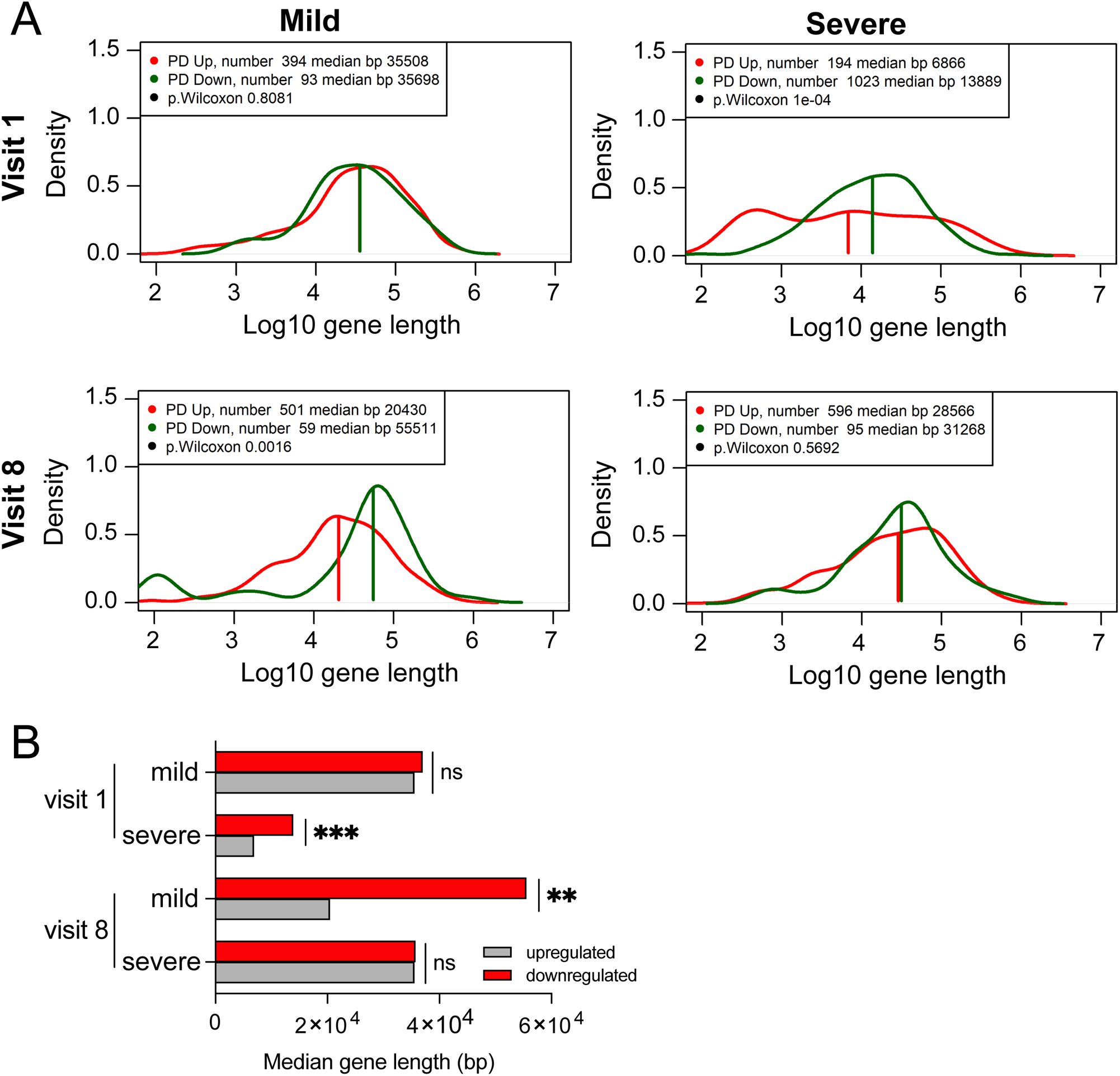
Biased reduction in the expression of longer genes in patients with different disease progression. (**A**) Frequency plots of gene length of up- and downregulated genes (red and green trace respectively) in iPD with mild and severe progression at visit 1 and 8. (**B**) Bar graph summarizing the results shown in the frequency plots (p.Wilcoxon, p<0. 01, **, p<0.001, ***). Statistical significance of the distributions was tested with Wilcoxon signed-rank test. Gene length is in log10 scale.

### Findings in peripheral cells mirror alterations in PD brains

To provide further evidence supporting the relevance of our findings in peripheral blood cells to the pathophysiology of the central nervous system, we employed a bioinformatic workflow known as Functional Mapping and Association analysis (FUMA). (*35, 36*). This tool allowed us to explore associations between expression profile traits and cell specificity.

In essence, the tool disentangles the complexity of a transcriptome originated from a pooled analysis, which includes multiple cell types, by comparing these results with expression patterns obtained at the single-cell level. FUMA thereby informs on the representation of cell-specific molecular signatures within the pooled dataset. Ultimately, the approach yields a cross-tissue imputation of a transcriptional signature.

When interrogated on tissue specific molecular signatures, not surprisingly FUMA analysis indicated that the most prominent contribution to the profile of DEG is given by the whole blood signature. Of note, the analysis also highlighted the contribution of several brain regions, particularly the *putamen* and the basal ganglia (fig.7A). Central Nervous System more represented genes were subject to GO over-representation analysis, which highlighted the involvement of processes relevant for PD that included dopamine uptake and biosynthesis, and dopamine-mediated neurotransmission (fig.7B). These results support that the alterations in DNA damage and repair we documented in blood of PD patients may parallel defects at the level of the brain. To provide conclusive evidence on the involvement of DNA damage and repair in PD-related neurodegeneration, applied ALBATRO to an openly available dataset (GSE68719) characterizing the transcriptome of the *substantia nigra* of 29 PD patients against 44 neurologically normal controls (*37*). The analysis revealed biased reduction in longer transcripts levels as expected (fig. S3D), therefore confirming the relevance of our findings in blood for the brain. We finally performed immunohistochemical (IHC) detection of the established DNA damage marker gammaH2AX in PD patients’ post-mortem tissues (PD N=9, healthy subjects N=7, demographics in fig. S3E). IHC in the *substantia nigra pars compacta* revealed a significant increase in signal in tyrosine hydroxylase positive neurons, when compared to controls (fig. 7C). These findings are fully consistent with the evidence we gained at bioinformatic level and, together we the data in peripheral cells we previously published (*16*), strongly implicate defective DNA repair and DNA damage accumulation in PD pathogenesis.

**Fig. 7.**
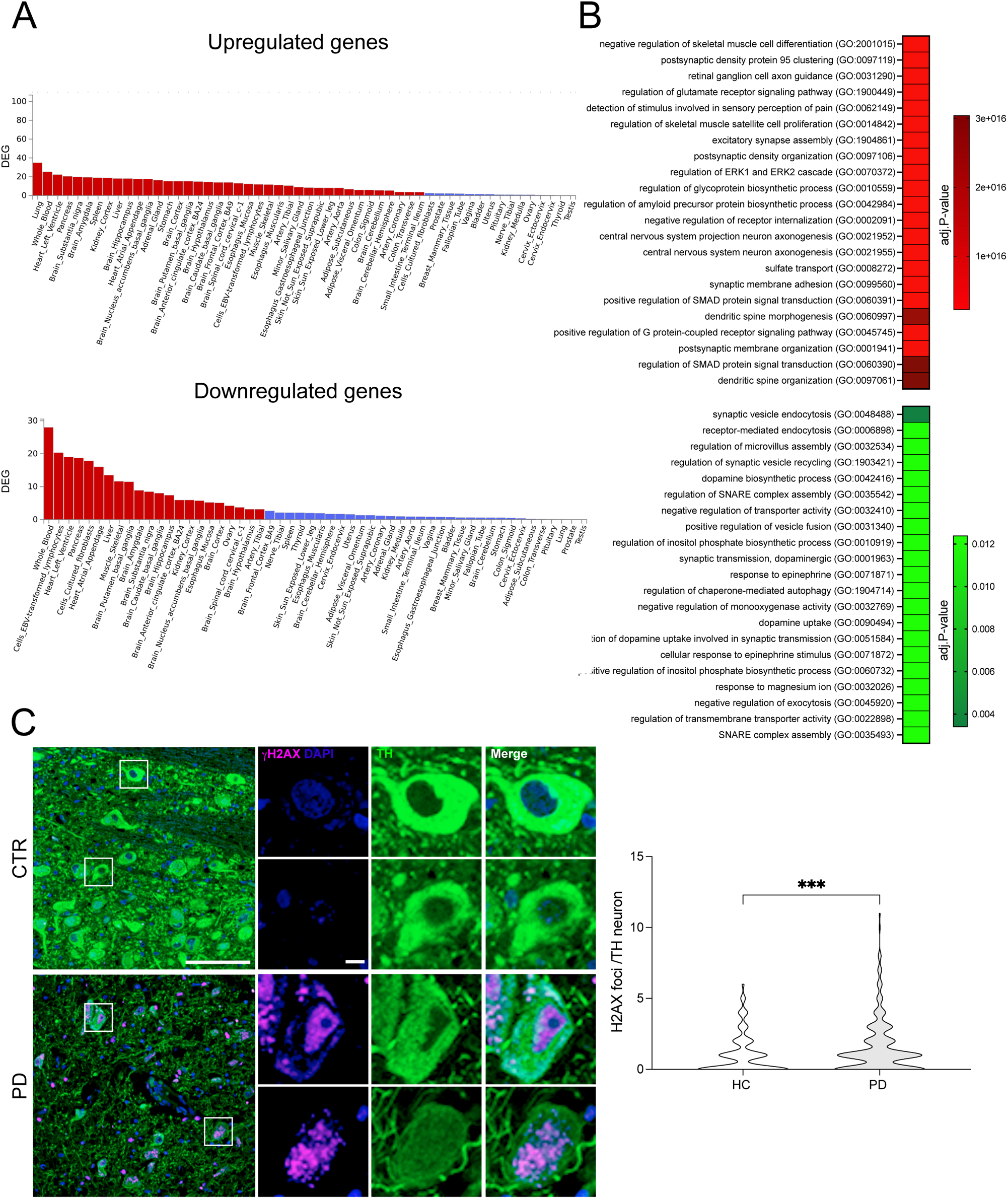
Functional mapping association analysis of statistically DEG in iPD samples. (**A**) The analytical framework indicates that the molecular signature includes a strong component of both up- and downregulated transcripts that are typical of cells in the putamen and basal ganglia brain regions. (B) Histogram showing the most representative up- and down-regulated genes in iPD blood that are also typical of cells in the *substantia nigra*. (**C**) Representative immunohistochemistry images of tyrosine hydroxylase positive neurons *substantia nigra pars compacta* of iPD patients (PD N=9, healthy subjects N=7) displaying gamma-H2AX foci (arrows). **(D)** Quantification of gamma-H2AX foci demonstrates increased signs of DNA damage signs iPD brains (p<0.0001, two-sided unpaired Student’s t-test).

## Discussion

Aging is PD principal risk factor and progressive accumulation of nuclear DNA damage is a causative hallmark of aging (*10*). In this study, we aimed to expand on previous evidence linking DNA damage and repair to PD by conducting a bioinformatic analysis of the blood transcriptome in the state-of-the-art longitudinal PPMI cohort. We confirmed our findings by analyzing independent cohorts and by performing immunochemical studies in autoptic PD brain tissues. While the data in genetic PD cases might be biased by reduced sample side, evidence in iPD is robust given that it is based on large experimental groups at both time points. The observed effects in patients are attributable to the pathogenesis and are not related to differences in age, given that the latter was comparable between PD patients and healthy subjects. Prodromal cases, however, were significantly older than healthy subjects and it is therefore possible that this difference is responsible for the observed alterations in this experimental group. Further studies will be necessary to conclusively confirm data in prodromal cases.

Our results revealed a reduction in the expression of DNA repair pathways and a distinct pattern of DNA damage accumulation in PD. These alterations were not caused by treatments, as indicated by analyses correcting for LEDD and medication types. Notably, the derangement of DNA repair pathways was primarily observed in patients who exhibited a faster progression rate over a three-year period. The reduced expression of DNA repair pathways preceded the biased reduction of long gene expression, which serves as a reliable surrogate marker for DNA damage accumulation (*26, 31, 32*). It is worth noting that the detection of DNA chemical modifications is challenging due to the wide variety of possible modifications, their rapid kinetics, and their diverse effects on cellular function (*38, 39*). These factors may lead to artefactual measures and consequently significant attention has been devoted to the use of alternative systems, for instance based on measures of repair efficiency (*40*). Biased reduction of long gene expression circumvents these limitations providing a measure of age-related DNA damage accumulation (*26*).

Recent studies have identified potential biomarkers, such as the α-synuclein seed amplification assay performed on cerebrospinal fluid – which is therefore invasive - (*41*) and a blood transcriptome signature (*42*). The distinctive strength of our signature lies in its ability to predict disease severity, particularly the progression of PD cardinal motor symptoms. This characteristic is highly relevant from the clinical standpoint because cardinal signs are the main targets of commonly used dopaminergic therapies and underlie the deterioration typical of the disease. Dopaminergic medications, however, have intrinsic complications that have a significant impact on patients’ quality of life and therefore require proper management (*43*). Our signature therefore lays foundation for better therapy management and patients’ wellbeing.

Our data highlight a remarkable difference in the transcriptomic landscape of slow versus fast progressors and indicate that, in the latter group, the transcriptome drifts significantly in the three-year period of observation. It is tempting to speculate reduced genome maintenance, which is observed in fast progressing patients, contributes to the evolution of the transcriptome. This phenomenon parallels what is observed in cancer (*44*), although genetic and transcriptional evolution in PD likely occur at a slower pace. Indeed, studying mouse models of genome instability we found that mild, rather than strong DNA repair defects elicit a dopaminergic phenotype recapitulating PD, also because of a pronounced adaptive, antioxidant response elicited by severe transcription stalling DNA damage (*16, 32*).

We did not detect any alterations related to DNA repair pathways in patients with GBA mutations, and the statistical power of our cohort may have limited the detection of significant alterations in LRRK2 mutants at the initial visit. However, it is possible that alterations related to DNA repair could have been detected at a later visit with a larger sample size. The absence of DNA repair alterations in the GBA cohort suggests distinct pathogenic mechanisms in this specific group of PD patients, which is consistent with the reported differences in disease presentation and age of onset among GBA carriers (*45*).

Stratification of patients is crucial in the field of neurodegenerative diseases (*46*). The heterogeneity of patients can in fact be fatal for the successful outcome of clinical trials because therapeutic interventions may have different effects depending on the individual pathogenic mechanisms driving disease progression. Our study provides evidence that defective DNA repair pathways can serve as predictive markers for the severity of motor symptoms and disease progression rate. Implementing this screening during patient recruitment for future trials could help refine patient selection, reduce heterogeneity in study cohorts, and design more informative clinical trials (*6, 8*).

Defects in genome maintenance have been hypothesized, investigated, and observed in several neurodegenerative disorders (*11, 47, 48*). Evidence gained in PD, in patients, is however limited (*49*), particularly at the level of large longitudinal cohorts. Our study strengthens the involvement of age-related DNA damage and repair in PD, consistent with previous findings from our laboratories demonstrating reduced DNA repair capacity in PD patients’ fibroblasts and impaired dopaminergic systems in DNA repair-deficient mice (*16*). These findings complement existing evidence indicating that alpha-synuclein activates the DNA damage response *in vivo* (*17*) and highlight the role of DNA damage accumulation in mitochondrial DNA in PD (*50, 51*).

Our findings in peripheral tissues have implications for the central nervous system. FUMA analysis, in fact, revealed that transcripts that are critical for brain functioning significantly contribute to the list of DEG. Furthermore, neuropathological analysis in post-mortem brains detected increased markers of DNA damage in PD. Overall, our study substantiates the concept that peripheral tissues can provide informative insights into central pathology and aligns with evidence showing that risk loci for PD impact various cellular processes, not necessarily confined to the brain, to one of its anatomical areas, or to a specific cell type (*52*).

DNA occupies the highest hierarchical position in biological information, and any compromise in its fidelity has broad impacts. DNA damage accumulation has been in fact associated with key mechanisms of neurodegeneration including mitochondrial defects, oxidant stress, proteotoxic stress, and inflammation (*53–57*). It is therefore plausible that defective DNA maintenance in PD interacts with other well-established pathogenic mechanisms. For instance, reduced nucleotide excision repair capacity sensitizes cells to MPTP and triggers alpha-synuclein stress, and LRRK2 biology impacts mitochondrial DNA quality (*16, 58*). Further focused studies are necessary to explore the mutual interactions and potential synergies between genome stability and PD pathogenic mechanisms.

Our study also presents some limitations that have to be addressed in future, dedicated investigations. For instance, the population of blood cells undergoes alterations in PD (*59*) and there is accelerated hemopoiesis during disease progression (*60*). These elements could influence the analyses we performed and differences in DNA repair may to some extent reverberate cellular changes in blood intrinsically associated with PD. This limitation could be circumvented, at least in part, taking advantage of deconvolution methods (*61*). A further limitation of our study may stem from the reduced size of patients harboring LRRK2 mutations, which might be responsible for the lack of an observed DNA repair phenotype. Indeed, this evidence contradicts other data from our and other laboratories indicating an effect of LRRK2 mutations on DNA damage and repair (*16, 58, 62*) and studies in cohorts including a larger number of LRRK2 patients are necessary to reach conclusive evidence. The same applies, to same extent, to GBA patients, which constituted groups of relatively small size in the analyzed PPMI cohort.

The main scope of this study was to identify a fingerprint of altered DNA maintenance in the PPMI cohort. We successfully detected these alterations in patients who exhibited a faster progression of motor symptoms over a 36-month observation period. Defects in DNA repair pathways may thus serve as predictive criteria for disease progression rate. Furthermore, our analysis revealed differences in the transcriptional landscape between slow and fast progressors, shedding light on potential variations in underlying pathogenic mechanisms. Collectively, our study identifies a biomarker predicting PD severity, reveals new pathogenic mechanisms, and demonstrates an evolution of PD blood transcriptome during disease progression. While further validation in other independent cohorts will be necessary to achieve conclusive validation and bring our findings to clinical practice, our results lay foundation for future tools to predict PD progression, improve therapy planning, and stratify patients in clinical trials.

## Materials and Methods

### Data and method

Blood transcriptome data from PPMI (Parkinson progressive markers initiative cohort) were downloaded at https://ida.loni.usc.edu/pages/access/geneticData.jsp#441. Briefly, data were processed in the following steps: the whole-transcriptome RNA-seq was obtained starting from 1 ug aliquots of RNA isolated from PaxGene tubes. Data RNA was sequenced at Hudson Alpha’s Genomic Services Lab on an Illumina NovaSeq6000. The samples were prepared using the NEB/Kapa (NEBKAP) based library prep. BCL’s were converted to using bcltofastq v1.8.4, and FASTQ’s were merged and aligned to GRCh38p12by STAR (v2.6.1d) on GENCODE v29. The IR3 (B38) (Phase1 and Phase2 count) was used for this analysis.

### Transcriptomic data preprocessing, quality control and batch correction

Downloaded Salmon files were imported into R using Tximport. Only transcripts with at least 10 reads in total were retained for analysis. Raw read counts were supplied to DESeq2, which was used to perform a differential expression analysis across PD groups and controls. The quality of the data was measured by identify the distribution of the gene counts, the library size, the clustering in a PCA, and finally in the ability to identify expected DEG between females and males.

To adjust for medication effects, we included medication variables as covariates in the design in DESeq2. The daily levodopa equivalent dose (LED) was included as an independent variable so that the model could account for their potential influence on the observed GSEA pathways. Medication type was treated as a categorical variable. In particular, we stratified patients into seven classes depending on the taken medications: (1) only levodopa/carbidopa, (2) only dopamine agonists, (3) levodopa/carbidopa and dopamine agonists, (4) levodopa/carbidopa and other medications, (5) levodopa/carbidopa, dopamine agonists, and other medications, (6) other medications, (7) no medications. When the medication filed was left blank in the PPMI record, the patient was classified as not taking any medication. No patients took dopamine agonists in combination with other medications. Other medications included A2A receptor antagonista, MAOB inhibitors, COMT inhibitors, and anticholinergics.

Rlog transformation was applied to the normalized counts to improve the distances/clustering for the PCA. To identify low quality samples, we performed visual inspection of the distribution of the reads.

### Identification of differentially expressed genes (DEGs)

We focused our investigations on two time points, that is at baseline (i.e. very first visit) and at visit 8 (i.e. 36 months after the first visit). Differential expressed genes (DEG) were estimated using log2 fold-change, the Wald test, and FDR p-value correction calculated as implemented in DESeq2. A gene was defined as differentially expressed (DEG) when its FDR was lower than 0.05.

### Identification of genetic variants in PD patients

Subjects with PD and controls and known genetic mutations associated with PD, such as GBA, LRRK2, and SNCA were enrolled in the genetic cohort and all the information can be found on www.ppmi-info.org. For this study we selected only subjects with a diagnosis of Parkinson’s disease and non-affected control subjects, with and without mutations in LRRK2_G2019S_ (LRRK2 patients) or in the GBA gene with the following mutations: GBA_N370S_, GBA_T408M_, GBA_E365K_, GBA_IVS2_, GBA_84GG_ or GBA_L444P_ (GBA patients). Demographic data are listed in table1.

### Pathway Analysis

Pathway enrichment analysis was conducted using two different approaches, overrepresentation analysis (ORA) (*23*) and gene set enrichment analysis (GSEA) (*24*). ORA focuses on significantly DEG (FDR ≤ 0.05) to inform whether these genes overrepresent known biological functions or processes. Because ORA processes only significantly DEG, it returns information based on relatively large differences. Functional differences, however, could stem from small, coordinated changes in group of related genes, i.e. expression modules, which by construction cannot be detected by a method bound to significantly DEG. GSEA addresses this limitation by analyzing all the genes ranked according the differences in expression between two groups.

ORA was performed via the EnrichR web tool using Gene Ontology 2018 (GO) for biological processes(*63–65*) from the pre-filtered list of differentially expressed genes obtained as described in the previous section. Fisher exact test was performed to determine the likelihood of obtaining at least the equivalent numbers of genes by as actually overlap between the input gene set and the genes present in each identified pathway.

GSEA was conducted on an unfiltered, ranked list of genes. Genes in each PD group compared with controls were ranked by the level of differential expression using a signal to noise metric, and a weighted enrichment statistic. Statistical significance of pathway enrichment score was ascertained by permutation testing over size matched random gene sets. Multiple testing was controlled by false positives with family-wise error rate (FWER) threshold of 5% (*24*), which is statistically more conservative than FDR.

We used GSEA v.4.2.2 that also includes the Kyoto Encyclopedia of Genes and Genomes (KEGG), Reactome Pathway Databases, Hallmark Gene Set Collection and WikiPathways (http://www.gsea-msigdb.org/gsea/msigdb/collections.jsp).

Pathway information was obtained from the Kyoto Encyclopedia of Genes and Genomes (KEGG) available at the Molecular Signatures Database (http://www.broadinstitute.org/gsea/msigdb/index.jsp) or from the Hallmark Gene Set Collection (http://www.gsea-msigdb.org/gsea/msigdb/collections.jsp).

### Gene Length analysis

BiomaRt (*66*) was used to retrieve the gene length (exons and introns) of all genes in the human genome. Each list of DEGs was divided in downregulated and upregulated genes. The distribution of the log10 length of up and down were evaluated by normality with a Shapiro–Wilk test. Because all the distributions were not normal, a Mann–Whitney Wilcoxon test for non-paired samples test was used to evaluate whether the distributions of gene lengths of up and down regulated DEGs were different between the different comparisons. Additionally, a relative frequency (kernel density) plot of gene length and probability density for DEG in each comparison was drawn using the density function implemented in R. Kernel density estimates are related to histograms, but with the possibility to smooth and continuity by using a kernel function. The y axis represents the density probability for a specific range of values in the x axis.

### Delta UPDRS classification of patients

Delta UPDRS was calculated subtracting UPDRS III (MDS-Unified Parkinson’s Disease Rating Scale) of last visit to the one of the first visit.

### Gene expression analysis with FUMA

FUMA is an online tool which identified significantly up- or downregulated differentially expressed gene sets across human tissue types with a Bonferroni corrected P < 0.05. GENE2FUNC, a tool of FUMA (https://fuma.ctglab.nl/) was run using GTex v8:tissue types and general tissue types (*35*).

An enrichment analysis of differentially tissue expressed genes (DEG) was carried put. The direction of expression was considered with <0,1. Tissue specificity is tested using hypergeometric tests. FUMA reports gene sets with adjusted P-value ≤ 0.05 and the number of genes that overlap with the gene set > 1 by default. Data are reported in a histogram representing the −log10 (p-value) with red (significant) and blue (not-significant) bars.

### Immunostaining procedures

Formalin fixed paraffin embedded fully anonymous human midbrain tissue sections derived from PD patients and age-matched healthy controls were kindly provided by the Queen Square Brain Bank for Neurological Disorders.

In order to minimize autofluorescence, sections have been treated for photobleaching as previously described (*67*). Briefly, samples were incubated in alkaline hydrogen peroxide solution (25mL PBS, 4.5 mL H_2_O_2_ 30%, 0.8mL NaOH 1M) in clear plastic petri dishes and exposed to white light by sandwiching the immersed slides between two light-emitting diode (LED) panels for 45’ at 4C (*67, 68*) for 45’ at 4C. The procedure has been repeated twice, with full solution change between the processes. Sections were then rinsed in TBS, pH 7.5, containing 0.01% Tween-20 and 100 mM sucrose (TBS-Ts) and blocked for non-specific binding with 5% donkey serum in antibody dilution buffer (ADB – 2% BSA, threalose 1 mM in TBS). Anti -tyrosine Hydroxylase (TH – 1:500, MAB 318, Merk Millipore) and anti -γH2AX (1:500 – ab11174, Abcam) have been used diluted in ADB overnight (ON) at 4C. After washes in TBS-Ts, sections have been incubated with alexa-fluor antibodies (1:250 in ADB, ThermoFisher) and DAPI (1:3000 in TBS-Ts). Finally, prolong Glass anti-fade medium (P36980, ThermoFisher) has been used as mounting medium. Images were acquired with the Leica SP8 STED imaging acquisition system at 40X magnification and gammaH2AX foci count was performed in a semi-automatic way using FIJI/imageJ software.

## Data Availability

All data produced in the present study are available upon reasonable request to the authors

https://ida.loni.usc.edu/pages/access/geneticData.jsp#441

## Acknowledgements

This study was supported by the Michael J Fox Foundation (PGM, JV). DS is supported by the Fondazione Veronesi.

