## Supplementary Figures for "Parkinson’s disease patients display a DNA damage signature in blood that is predictive of disease progression"

A

| Group | Visit 1 | | | Visit 8 | | |
|---|---|---|---|---|---|---|
| | Total | Male | % Male | Total | Male | % Male |
| iPD | 242 | 162 | 67 | 206 | 139 | 67 |
| GBA | 50 | 25 | 50 | 28 | 15 | 54 |
| LRRK2 G2019S | 134 | 66 | 49 | 14 | 6 | 43 |
| Prodromal | 58 | 46 | 79 | | | |
| Healthy subjects | 187 | 122 | 65 | 157 | 97 | 62 |

B

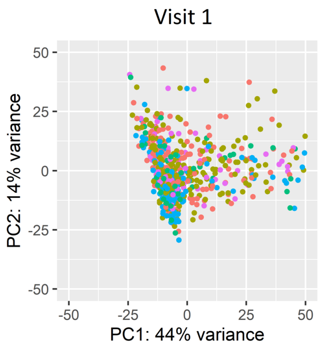


C

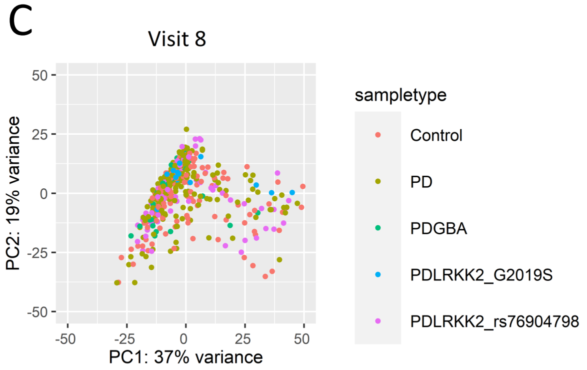


D

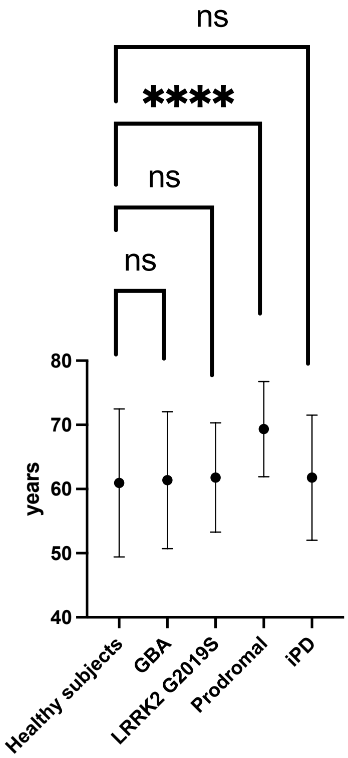


| | Age (mean ± SD) |
|---|---|
| Control | 60.94069±11.538212 |
| GBA | 61.38085±10.668944 |
| LRKK2_G2019 | 61.79847± 8.525879 |
| PD | 61.77045± 9.745930 |
| Prodromal | 69.33529± 7.424027 |

E

| | Total DEG | Common DEG | EF |
|---|---|---|---|
| Visit 1 | 237 | 69 | 26.92 |
| Visit8 | 216 | | |

Supplementary fig.1

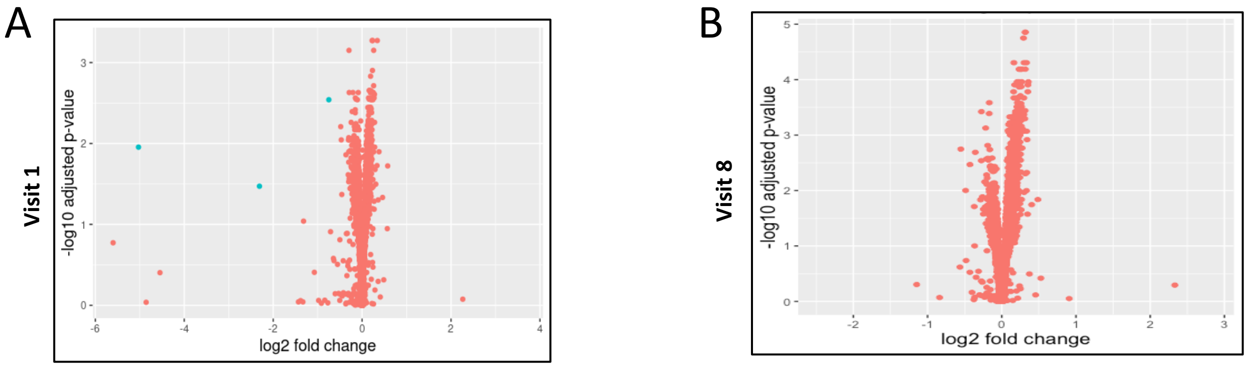


C

| Number of DE genes, Log2FC>0.1 or Log2FC<-0.1, padj<0.05 | | | | |
|---|---|---|---|---|
| | Visit1 | | Visit8 | |
| | UP | DOWN | UP | DOWN |
| iPD | 83 | 70 | 628 | 54 |
| GBA | 114 | 74 | 580 | 1590 |
| LRRK2 G2019S | 8538 | 2419 | 28 | 41 |
| Prodromal | 7880 | 3797 | | |

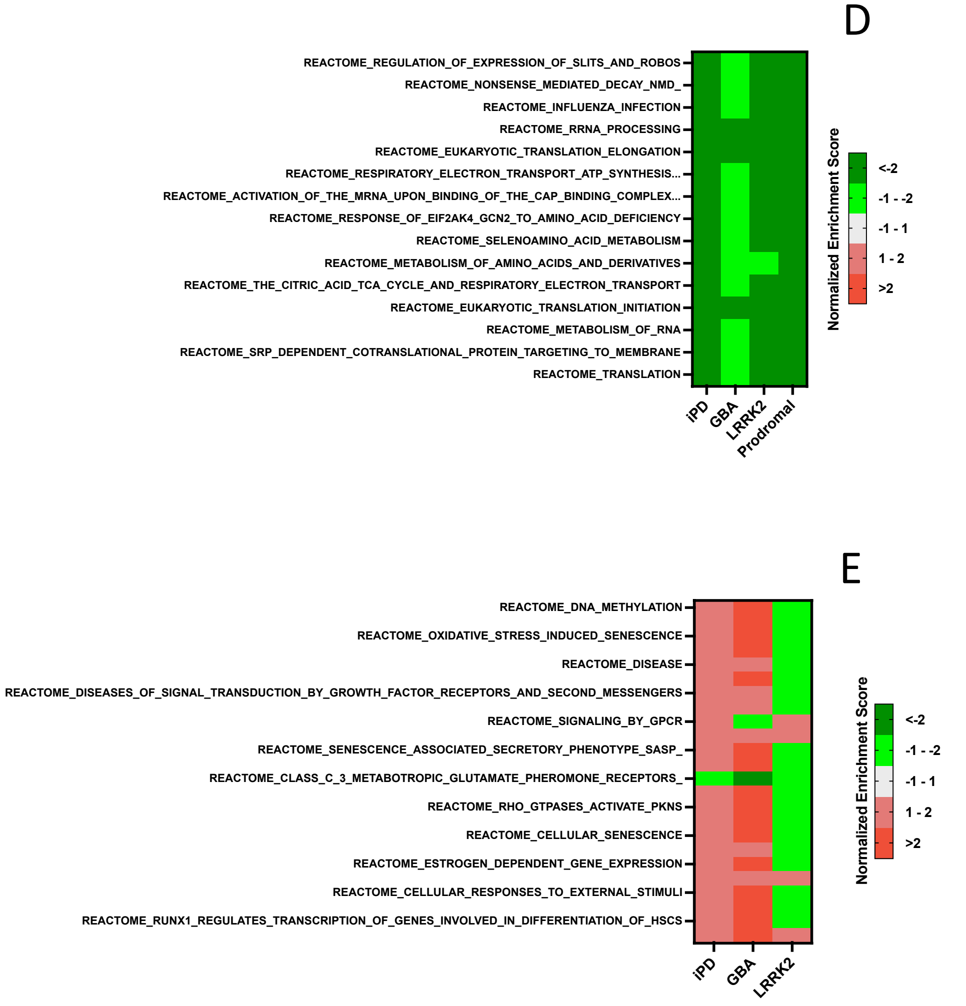


Supplementary fig.2

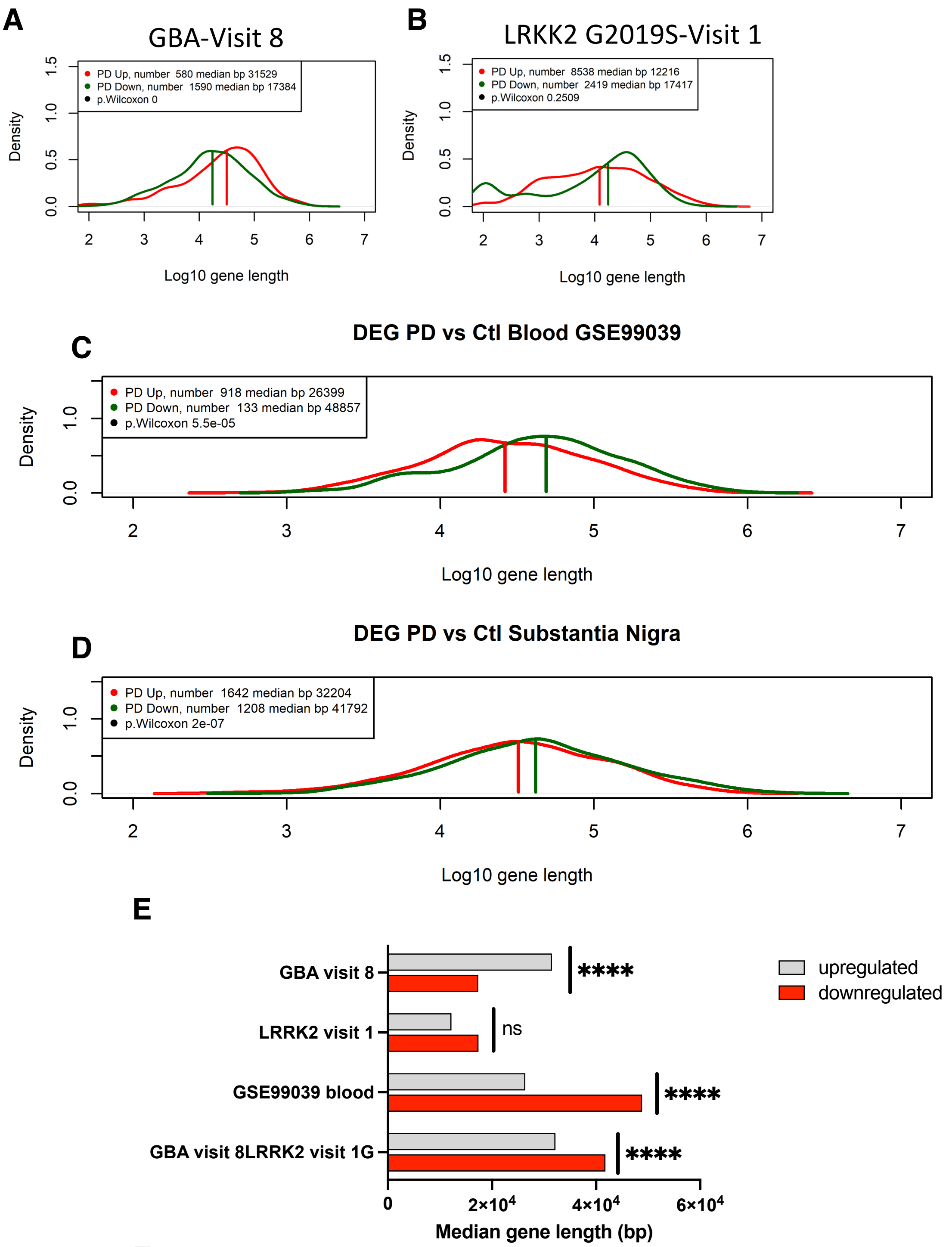


**F**

| Group | Age at onset (Average +/- SD) | Age at death (Average +/- SD) | Gender (M/F) |
|---|---|---|---|
| **IPD** | 59.4 +/- 4.9 | 78.3 +/- 3.7 | 4/5 |
| **Control** | NA | 77.7 +/- 10 | 4/3 |

Supplementary fig.3

**Supplementary figures legends**

**Fig. S1. PCA and volcano plots of PD, Genetic cohort, Prodromal and healthy controls at visit number 1 and visit number 8.** (**A**) Demographic tables summarizing number and sex of patients in the four PD subgroups and healthy controls. (**B**, **C**) Principal component analysis plots. (**D**) Aging of PD patients' groups is comparable with that of healthy subjects, as indicated in the graph (right) and in the table (left). Prodromal patients, however, are significantly older than healthy subjects (**** P<0.0001, Brown-Forsythe and Welch ANOVA test). (**E**) Table illustrating the enrichment factor of common deregulated genes at visit 1 and visit 8.

**Fig. S2. Volcano plots of differentially expressed genes in iPD.** DEG at visit 1 (**A**) and at visit 8 (**B**). Blue dots are genes with Log2FC>0.584 or Log2FC<-0.584 and padj<0.05. Table C lists the number of DE genes with Log2FC>0.1 or Log2FC<-0.1, adj.P-value<0.05. (**E, F**) Heatmap showing expression changes in pathways common to the different experimental groups at visit 1 and 8.

**Fig. S3. Analysis for length-biased alterations in transcription output in additional datasets.** ALBATRO frequency plots of gene length in the GBA cohort at visit 8 (**A**), LRKK2-G2019S (**B**). (**C**) Frequency plots of gene length in a different PD blood dataset (GSE99309). (**D**) Frequency plots of gene length in a PD dataset from the *substantia nigra* (GSE68719). (**E**) Histogram summarizing the results in the frequency plots. Note how GBA patients behave in the opposite way than the iPD, displaying shorter downregulated genes (p.Wilcoxon, p<0.0001 ****). (**F**) Table describing the characteristics of the Queen Square Brain Bank (QSBB) *substantia nigra pars compacta* specimens.
